# Retention to Care and Viral Load Suppression: Insights from Young People Receiving HIV Treatment at Mpilo Centre of Excellence in Bulawayo, Zimbabwe

**DOI:** 10.64898/2026.03.28.26349591

**Authors:** Primrose S Dube, Sinatra Nyathi, Nkazimulo I Tshuma, Solwayo Ngwenya, Masimba Masiya, Douglas Moyo, Clearance Maruba, Freeman Dube, London Makwanya, Raymond Yekeye, Amon Mpofu, Bernard Madzima

## Abstract

**Background:** Retention to care and viral load suppression are essential components for effective HIV management, particularly among adolescents and young adults aged 15–24 years, who remain vulnerable to treatment challenges. This study aimed to assess factors associated with poor retention in care and viral load suppression among young people receiving antiretroviral therapy (ART) at Mpilo Centre of Excellence (MCoE) in Bulawayo, Zimbabwe, with the objective to guide youth-friendly interventions and improve health outcomes.

**Methods:** A mixed methods cross-sectional study was conducted involving 110 HIV-positive youths aged 15-24 years on ART, recruited through systematic sampling and surveyed between November and December 2024. Data was collected using structured questionnaires, focus group discussions, in-depth interviews, and key informant interviews. Quantitative data were analyzed using descriptive statistics and logistic regression models to identify factors linked to viral load suppression, while qualitative data underwent thematic analysis.

**Results:** Viral load suppression was achieved by 68.19% of participants, who met the viral suppression criterion of <50 copies/ml. Analysis identified several significant predictors via multivariable logistic regression. Younger adolescents (15–19 years) had lower odds of achieving suppression compared to older youths (20–24 years) (Adjusted Odds Ratio [AOR]: 0.81; 95% Confidence Interval [CI]: 0.67–0.97; p=0.041), while female participants demonstrated higher suppression rates than males (AOR: 0.43; 95% CI: 0.21–0.96; p=0.032). Absence of adherence challenges to ART emerged as a strong predictor of suppression (AOR: 0.12; 95% CI: 0.03–0.72; p=0.018), and perceived lack of clinical staff support was associated with a threefold higher risk of unsuppressed viral load (AOR: 3.01; 95% CI: 1.34–7.69; p=0.046). Lower treatment self-efficacy negatively impacted suppression odds (AOR: 2.65; 95% CI: 1.11–7.83; p=0.046), and lack of friend support for clinic visits reduced the likelihood of suppression (AOR: 0.31; 95% CI: 0.09–0.89; p=0.001). Qualitative findings confirmed that persistent barriers—including stigma, limited family support, economic hardship, school and work commitments—compromised both retention and adherence among adolescents and young adults.

**Conclusion:** Younger age, male sex, ART adherence challenges, lack of clinical staff support, and lower treatment self-efficacy were significantly associated with poor viral suppression among 15–24-year-olds at Mpilo Centre of Excellence. These findings underscore the need for tailored adolescent- and youth-friendly services, enhanced adherence support, and improved treatment literacy to strengthen retention in care and viral suppression. Addressing these factors is critical for advancing progress towards UNAIDS 95-95-95 targets and reducing HIV transmission among Zimbabwean youth.

## Introduction

Retention to care and viral load suppression are critical components in the successful management of HIV, especially among adolescents and young adults aged 15 to 24 years. Retention in care refers to the continuous engagement of patients with HIV treatment services and is essential for achieving and maintaining viral load suppression. Viral suppression not only improves individual health outcomes but also reduces onward HIV transmission, contributing significantly to epidemic control. This study aimed to assess the factors associated with poor retention to care and viral load suppression among 15–24-year-olds accessing services at the Mpilo Centre of Excellence (MCoE) in Bulawayo, Zimbabwe, a population particularly vulnerable to treatment challenges.

A growing body of literature highlights numerous interconnected determinants that compromise retention and viral suppression outcomes in young people living with HIV. Psychosocial challenges such as stigma, mental health issues, and lack of social support frequently disrupt adherence and clinic attendance [1], [2], [3]. Socioeconomic factors including poverty and transportation barriers further restrict consistent access to care [4]. Clinical factors like delayed viral load testing and co-infections also contribute to treatment failure [5]. Studies from MCoE and similar contexts report that only about half of adolescents achieve viral suppression six months after ART initiation, with attrition rates increasing with age within the adolescent and young adult group [6], [7]. Despite the implementation of differentiated service delivery programs such as Community Adolescent Treatment Supporters (CATS) and Family ART Refill Groups (FARGS), retention and viral suppression remain inadequate, signaling persistent gaps in program reach and effectiveness.

At MCoE, viral load suppression among 15–24-year-olds has consistently fallen short of national and global targets. Retention data reveal significant losses along the treatment cascade, threatening efforts to meet the UNAIDS 95-95-95 goals for this key population[8]. This study seeks to identify and analyze the factors associated with poor retention to care and viral load suppression within this demographic to inform the design of targeted, youth-friendly interventions. Addressing these complex determinants is essential to improving treatment outcomes and reducing HIV transmission among young people at Mpilo Centre of Excellence.

## Materials and Methods

### Study Design

The study employed a mixed methods cross sectional study design. We also incorporated a qualitative component to further enrich the study findings.

### Study Setting and Population

We conducted the study at Mpilo Centre of Excellence in Bulawayo, Zimbabwe. Eligible participants included HIV-positive recipients of ART aged 15 to 24 years who accessed care between January 2018 and May 2024 with documented HIV status disclosure.

### Variables

The following variables guided the research:

- **Outcomes:** Retention to care and viral load suppression among 15–24-year-olds on ART at Mpilo Centre of Excellence.
- **Exposures and predictors**: Demographic characteristics (age, sex, marital status, education, occupation, residence), treatment adherence factors (challenges in taking ART, friend support for adherence and clinic visits), clinical factors (perceived clinical staff support), behavioral and self-efficacy factors (confidence in taking ART as directed, belief in ART efficacy), and baseline clinical measures (years on ART).
- **Potential confounders/effect modifiers:** Age, sex, living arrangements and social support factors.
- **Diagnostic criteria:** Viral load suppression defined as <50 copies/ml; incident virologic failure as ≥50 copies/ml confirmed after prior suppression.

### Data Sources/Measurement

The study used structured questionnaires and interviews. Demographic, clinical, and treatment adherence data was collected via structured surveys. Standardized questionnaires and interview guides were used with qualitative and quantitative data triangulated for validation.

### Bias

Efforts to minimize bias included systematic sampling from the complete eligible population to reduce selection bias. The use of mixed methods (quantitative and qualitative) to triangulate data enhanced validity. Data collectors were trained and supervised to minimize information bias. Data validation checks and electronic data collection controls were used to reduce data entry errors which include skip logics. The team was guided by ethical safeguards and confidentiality which encouraged truthful reporting.

### Study Size

A sample size of 107 was calculated using Cochran formula, factoring attrition, based on comorbidity proportions from a Ugandan study.

### Sampling Strategy and Representativeness

The study employed systematic random sampling from a comprehensive registry of all HIV-positive adolescents and young adults aged 15 to 24 years accessing ART at Mpilo Centre of Excellence between January 2018 and May 2024. Eligibility required documented HIV status disclosure and at least one viral load measurement within the treatment period. The sample size of 110 was determined using Cochran’s formula, accounting for an anticipated attrition rate and prevalence of poor viral suppression from prior regional studies. Systematic sampling ensured proportional representation across age groups, sex, and geographical areas served by the clinic, enhancing the generalizability of findings to the broader youth population at Mpilo. Continuous monitoring of demographic characteristics during recruitment ensured no subgroup was underrepresented.

### Quantitative Variables

Age was grouped into 15-19 (teenagers) and 20-24 (young adults) to capture developmental differences. Viral load was treated as a binary categorical variable (suppressed <50 copies/ml vs unsuppressed ≥50 copies/ml). Occupation was categorized into student, unemployed, self-employed, part-time and full-time employed. Living arrangements were grouped into parental vs non-parental care. Continuous variables like age and years on ART were used both as continuous variables and categorical groupings for analyses. Logistic regression models were used to analyze associations, with variable inclusion based on p-value and clinical relevance.

### Data Collection

A structured questionnaire was administered to 227 youths to capture adherence, social support, and behavioral factors. Six focus group discussions (FGDs), 16 in-depth interviews (IDIs) with patients, and nine key informant interviews (KIIs) with healthcare workers and caregivers were conducted.

### Data Analysis

Quantitative data was analysed using Stata 17 and a summary of demographic and outcome data with descriptive statistics was produced. Factors associated with viral suppression were identified using univariable and multivariable logistic regression. Qualitative data was coded and thematic analysis performed using Dedoose software.

### Statistical methods

#### (a) Statistical Methods and Control for Confounding

Cross tabulations and statistical tests were conducted to interpret data. We first performed univariable analyses to identify potential variables associated with viral suppression and other outcomes. Variables with p-value < 0.25 in univariable analyses or considered clinically relevant were then included in multivariable logistic regression models. Multivariable logistic regression was used to estimate adjusted odds ratios (AORs) controlling for potential confounders such as age, sex, education level, living arrangements, treatment adherence challenges, friend support, and clinical staff support.

#### (b) Examination of Subgroups

Age was analyzed both as a continuous variable and in categorical groupings (15-19 years vs 20-24 years) to examine differences between adolescents and young adults. Gender subgroups were examined for their association with viral suppression and retention.

#### (c) Handling of Missing Data

Among the 227 youths initially approached for the cross-sectional study, 117 were excluded from viral suppression analyses due to incomplete or unavailable viral load data. To address this, we performed data validation checks prior to analysis. Missing viral load data were assessed for randomness; no significant demographic or clinical differences were observed between those with missing versus available viral load results, suggesting minimal systematic bias. Cases with missing viral load measurements were excluded from regression models focused on viral suppression outcomes. Sensitivity analyses were considered but limited due to lack of auxiliary information. Future studies are encouraged to strengthen routine viral load monitoring and data completeness to reduce missingness.

### Rationale for Mixed Methods and Data Integration

The mixed methods design was intentionally selected to comprehensively explore factors influencing retention and viral suppression among youths. Quantitative data provided measurable associations between demographic, clinical, and behavioral variables and viral suppression outcomes, allowing for statistical inference. Complementarily, qualitative data from focus groups, in-depth interviews, and key informant interviews elucidated contextual experiences, attitudes, and barriers that quantitative metrics alone could not capture. Integration occurred through triangulation, whereby emerging themes from qualitative analysis explained and enriched interpretation of quantitative findings. For example, quantitative associations with clinical staff support were deepened by qualitative accounts of patient-provider interactions, reinforcing the importance of a supportive clinic environment. This approach strengthened the validity and applicability of results, informing more tailored and effective youth-friendly intervention strategies.

### Ethical Considerations

All participants took part in the study only after providing informed consent or assent, as appropriate. Research participants aged 18 years and above provided their own written informed consent, while those younger than 18 years provided written assent, with parallel written informed consent obtained from their parents or legal guardians. In addition, informed consent was sought from partners and other stakeholders, including specific consent for audio recording, and no photographs were taken. Confidentiality and anonymity were strictly maintained throughout, with the Evaluation Team treating every response as confidential and not collecting participants’ names or other directly identifying information. No participant was excluded from participating on condition of their health, disability, socio-economic status, among other vulnerability criteria. Risks were continuously assessed with the research avoiding putting participants and the research team at risk. Ethics approval for the study was granted by the National AIDS Council Internal Review Board **(2024 NAC REC 01)**, and permission to use clinical data from Mpilo Centre of Excellence was obtained from the Centre. All procedures followed the principles of the Declaration of Helsinki (18th WMA General Assembly, Helsinki, 2001), ensuring respect for participants’ life, health, privacy and dignity, minimisation of harm, and that the anticipated benefits and importance of the research outweighed any potential risks.

## Results

### Respondents Flow Chart

Of the 2,342 eligible adolescents and young adults receiving antiretroviral therapy (ART) at Mpilo Centre of excellence, 227 participated in the study. Data collection was conducted between November 19 and December 7, 2024, using structured interviews. Of the enrolled ART recipients, 117 were excluded from the analysis due to incomplete or unavailable viral load data. As a result, 110 participants met the inclusion criteria and were included in the analysis for the study as shown in Figure 1.

**Figure 1:**
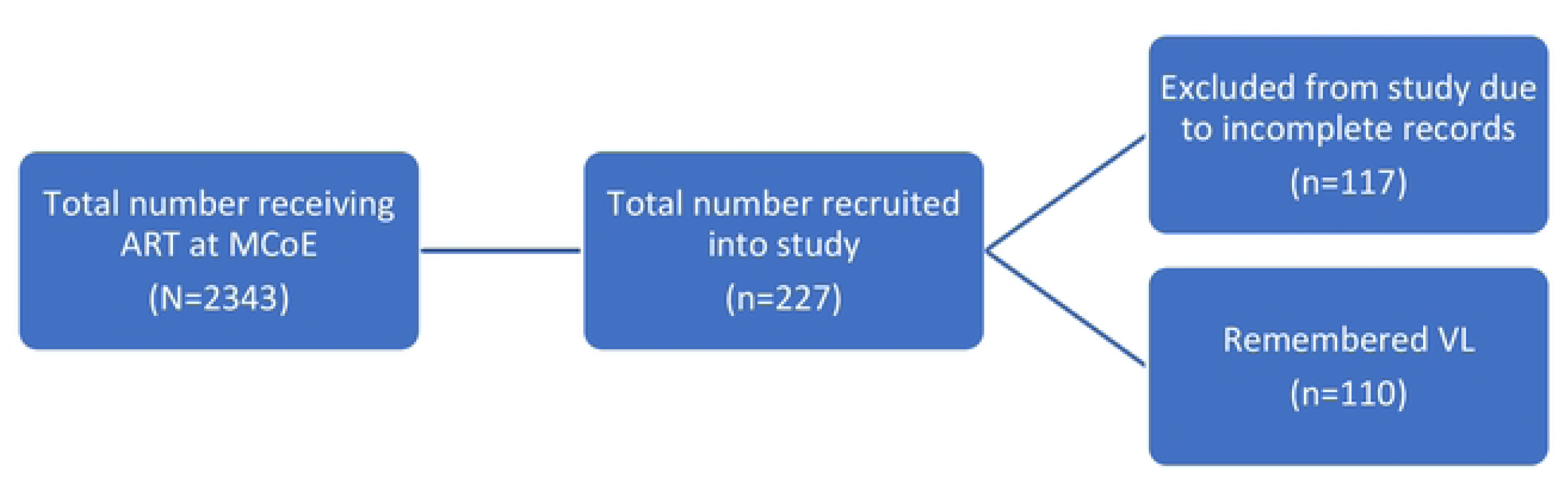
Respondents flow chat.

### Participant Characteristics (Remembered Viral Load)

**Table 1:** Demographic characteristics of participants who remembered their viral load.

|  |  |  |
| --- | --- | --- |
| **Age range** | 15 –19 years | n = 110<br>41 (37.27%) |
|  | 20 – 24 years | 69 (62.73%) |
| **Age (continuous)** | Mean (standard deviation) | 20.22 (2.66) |
|  | Median (IQR) | 20 |
| **Sex** |  | n = 110 |
|  | Male | 49 (44.55%) |
|  | Female | 61 (55.45%) |
| **Marital Status** | Single<br>Married | n = 110<br>105 (95.45%)<br>5 (4.55%) |
| **Level of Education** | Never gone to school<br>Primary<br>Secondary<br>Tertiary | n = 110<br>1 (0.91%)<br>5 (4.55%)<br>89 (80.91%)<br>15 (13.64%) |
| **Occupation** | Student<br>Unemployed<br>self-employed<br>employed part time<br>employed full time | n = 110<br>34 (30.91%)<br>44 (40.00%)<br>9 (8.18%)<br>11 (10.00%)<br>12 (10.91%) |
| **District of residence (MoHCC demarcation)** | Emakhandeni<br>Nkulumane<br>Northern<br>Out of Bulawayo | n = 110<br>43 (39.09%)<br>34 (30.91%)<br>32 (29.09%)<br>1 (0.91%) |
| **Who Participant Lives with** | Mother and Father<br>Mother<br>Father<br>other | n = 110<br>17 (15.45%)<br>30 (27.27%)<br>7 (6.36%)<br>56 (50.91%) |

Of the 110 recipients of care on antiretroviral therapy (ART) included in the cross-sectional analysis, 49 (44.55%) were male, and 61 (55.45%) were female. The mean age of participants was 20.22 years (median = 20 years). Age was categorized into two groups to differentiate between teenagers (15–19 years) and young adults (20–24 years) and to assess potential differences in viral load suppression. The majority of participants were single (n = 105, 95.45%), with only 5 (4.55%) being married. Nearly all participants had attained at least a primary school education (n = 109, 99.09%), and 104 (94.55%) had completed at least secondary school. In terms of occupation, 34 participants (30.91%) were students, 44 (40.00%) were unemployed, and 32 (29.09%) were employed.

### Factors associated with viral suppression

**Table 2:** Factors associated with viral load suppression.

| Demographic Characteristic | Viral Load |  | Odds Ratio | 95% Confidence Interval | Probability Value |
| --- | --- | --- | --- | --- | --- |
|  | High (>50/ml) | Low (<50/ml) |  |  |  |
| Age range | n = 35 | n = 75 |  |  |  |
| 15 – 19 years | 16 (45.71%) | 25 (33.33%) | 1 (ref) |  |  |
| 20 – 24 years | 19 (54.29%) | 50 (66.67%) | 0.59 | 0.26 – 1.34 | 0.213 |
| Age (continuous) | 35 (31.81%) | 75 (68.19%) | 0.85 | 0.73 – 0.99 | 0.042* |
| Sex | n = 35 | n = 75 |  |  |  |
| Male | 21 (60.00%) | 28 (37.33%) | 1 (ref) |  |  |
| Female | 14 (40.00%) | 47 (62.67%) | 0.40 | 0.17 -0.90 | 0.028* |
| Marital Status | n = 35 | n = 75 |  |  |  |
| Married | 1 (2.86%) | 4 (5.33%) | 0.52 |  |  |
| Single | 34 (97.26) | 71 (94.67%) | 1 (ref) | 0.05 – 4.85 | 0.561 |
| Level of Education | n = 35 | n = 75 |  |  |  |
| Never gone to school | 0 (0.00%) | 1 (1.33%) |  |  |  |
| Primary | 0 (0.00%) | 5 (6.67%) | - |  |  |
| Secondary | 30 (85.71%) | 59 (78.67%) | 1.02 |  |  |
| Tertiary | 5 (14.29%) | 10 (13.33%) | 1 (ref) | 00.32 - 3.24 | 0.32 - 3.24 |
| Occupation | n = 35 | n = 75 |  |  |  |
| Student | 17 (48.57%) | 17 (22.67%) | 1 (ref) |  |  |
| unemployed | 8 (22.86%) | 36 (48.00%) | 0.22 | 0.08 - 0.62 | 0.004* |
| self-employed | 3 (8.57%) | 6 (8.00%) | 0.5 | 0.11 - 2.33 | 0.378 |
| employed part time | 3 (8.57%) | 8 (10.67%) | 0.38 | 0.08 - 1.66 | 0.196 |
| employed full time | 4 (11.43%) | 8 (10.67%) | 0.5 | 0.13 - 1.98 | 0.323 |
| Who Participant Lives with | n = 35 | n = 75 |  |  |  |
| Mother and Father | 9 (25.71%) | 8 (10.67%) | 1 (ref) |  |  |
| Mother | 8 (22.86%) | 22 (29.33%) | 0.32 | 0.09 - 1.13 | 0.077* |
| Father | 1 (2.86%) | 6 (8.00%) | 0.15 | 0.01 - 1.51 | 0.107 |
| other | 17 (48.57%) | 39 (52.00%) | 0.39 | 0.12 - 1.18 | 0.094* |

Age was analysed both as a continuous and categorical variable to evaluate whether its association with viral suppression varied by coding. Age demonstrated a statistically significant association with viral suppression when treated as a continuous variable (OR 0.85; p=0.042). Sex was also significantly associated with viral suppression, indicating differences in suppression status between males (OR 0.40; p=0.028) and females (OR 0.43; p=0.032), with women likely to be at greater risk. Additionally, level of education and living arrangements had p-values < 0.25, but not statistically significant suggesting potential relevance for inclusion in the multivariable logistic regression model. Other variables not shown in the table had p values that were greater than 0.3.

**Table 3:** Treatment adherence factors.

|  |  |
| --- | --- |
| Treatment Adherence Factors | Viral Load |

|  | High<br>(>50/ml) | Low<br>(<50/ml) | Odds<br>Ratio | 95%<br>Confidence<br>Interval | Probability<br>Value |
| --- | --- | --- | --- | --- | --- |
| Challenges in taking ART | n = 35 | n = 75 |  |  |  |
| No | 1 (2.86%) | 15 (20%) | 1 (ref) |  |  |
| Yes | 34 (97.14%) | 60 (80.00%) | 0.11 | 0.01 – 0.93 | 0.042* |
| Have friend support in ART | n = 35 | n = 75 |  |  |  |
| No | 29 (82.86%) | 47 (62.67%) | 2.87 | 1.06 – 7.79 | 0.037* |
| Yes | 6 (17.14%) | 28 (37.33%) | 1 (ref) |  |  |
| Friend support in clinical appointments | n = 35 | n = 75 |  |  |  |
| No | 31 (88.57%) | 56 (74.67%) | 2.63 | 0.82 – 8.42 | 0.104 |
| Yes | 4 (11.43%) | 19 (25.33%) | 1 (ref) |  |  |

Participants who reported experiencing challenges in taking antiretroviral therapy (ART) during the preceding three months exhibited differences in viral suppression compared to those who did not face any difficulties in ART adherence. Additionally, participants who received support from friends in adhering to ART showed a distinct viral suppression status compared to those without such support (OR 2.87; p=0.016)

**Table 4:**
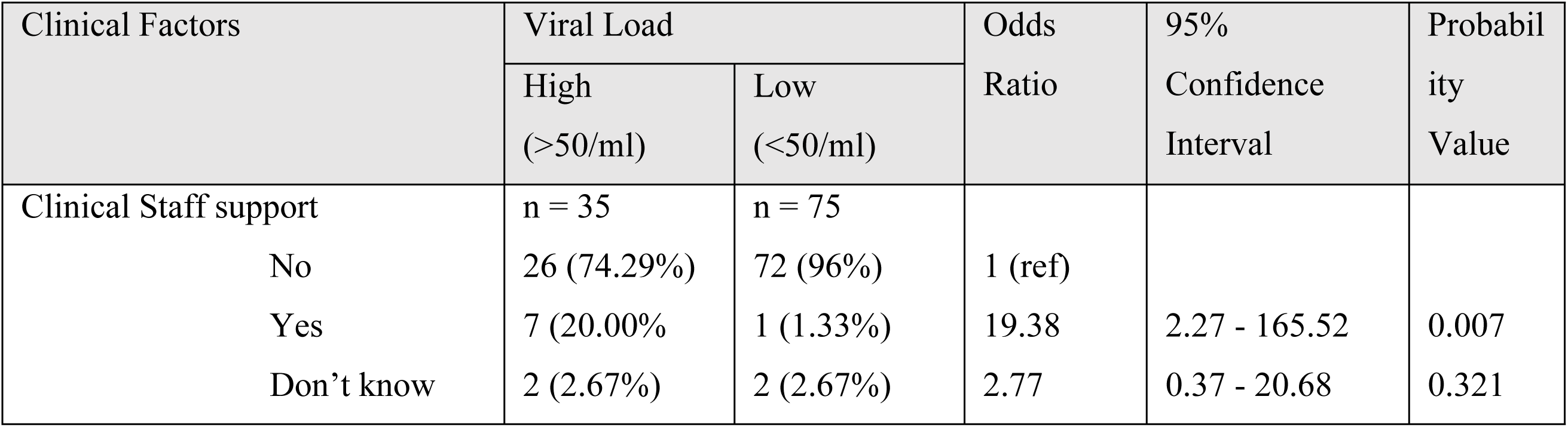
Clinical factors.

| Clinical Factors | Viral Load |  | Odds<br>Ratio | 95%<br>Confidence<br>Interval | Probabil<br>ity<br>Value |
| --- | --- | --- | --- | --- | --- |
|  | High<br>(>50/ml) | Low<br>(<50/ml) |  |  |  |
| Clinical Staff support | n = 35 | n = 75 |  |  |  |
| No | 26 (74.29%) | 72 (96%) | 1 (ref) |  |  |
| Yes | 7 (20.00%) | 1 (1.33%) | 19.38 | 2.27 - 165.52 | 0.007 |
| Don't know | 2 (2.67%) | 2 (2.67%) | 2.77 | 0.37 - 20.68 | 0.321 |

The only statistically significant factor was whether or not the participant felt that clinical staff were supportive. Those that felt so, had a different viral suppression status to those that felt clinical staff were not supportive. All other clinical factors were not statistically significant.

**Table 5:**
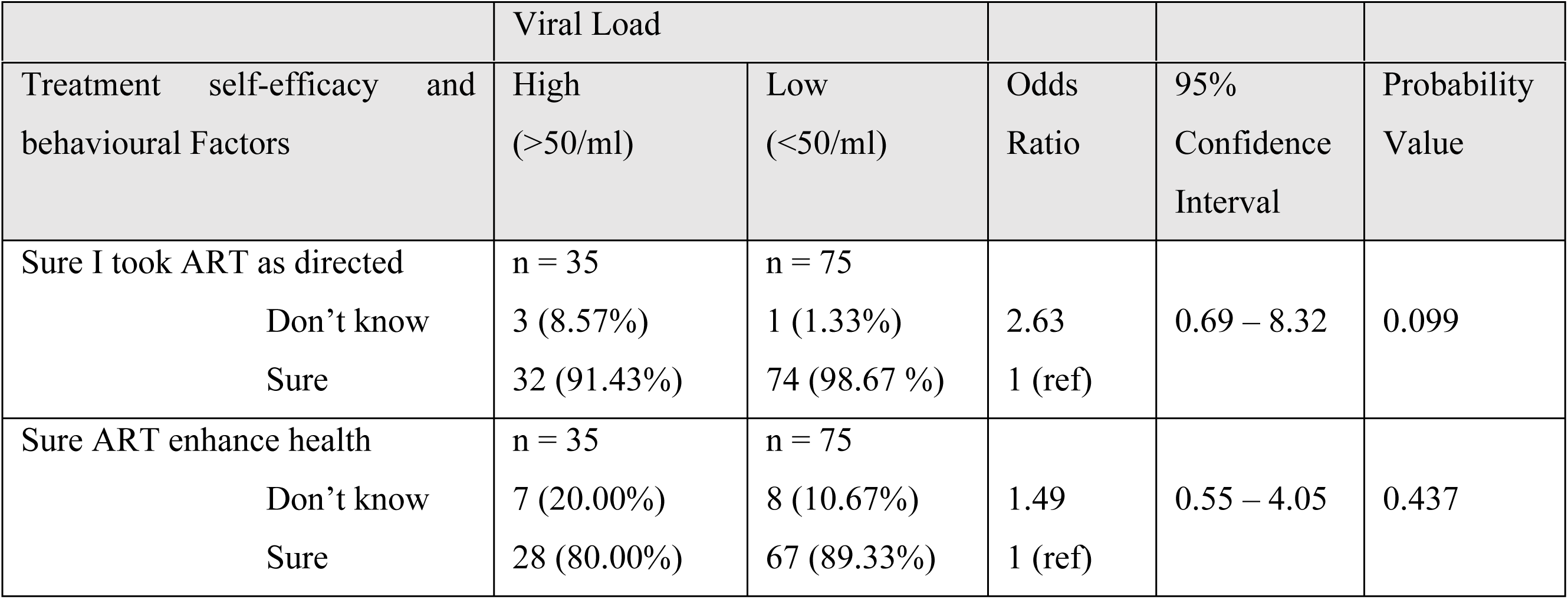
Univariable Analysis of treatment self-efficacy and behavioural factors.

| Treatment self-efficacy and behavioural Factors | High (>50/ml) | Low (<50/ml) | Odds Ratio | 95% Confidence Interval | Probability Value |
| --- | --- | --- | --- | --- | --- |
| Sure I took ART as directed | n = 35 | n = 75 |  |  |  |
| Don't know | 3 (8.57%) | 1 (1.33%) | 2.63 | 0.69 – 8.32 | 0.099 |
| Sure | 32 (91.43%) | 74 (98.67 %) | 1 (ref) |  |  |
| Sure ART enhance health | n = 35 | n = 75 |  |  |  |
| Don't know | 7 (20.00%) | 8 (10.67%) | 1.49 | 0.55 – 4.05 | 0.437 |
| Sure | 28 (80.00%) | 67 (89.33%) | 1 (ref) |  |  |

The table above presents the univariable analysis of treatment self-efficacy and behavioral factors associated with viral suppression. Among these factors, only one demonstrated statistical significance: Sure I took ART as directed. Additionally, two factors with p-values below 0.25 were identified as candidates for multivariable modelling. These factors were: a participant’s confidence in taking ART as directed and their belief that ART enhances health and prolongs life.

**Table 6:** Univariable and Multivariable Logistic regression for Viral suppression.

| Variable | Crude Odds Ratio (CI) | p-value | Adjusted Odds Ratio (CI) | p-value |
| --- | --- | --- | --- | --- |
| Age (continuous) | 0.85 (0.73 – 0.99) | 0.042 | 0.81 (0.67 – 0.97) | 0.041 |
| Sex (male) | 0.40 (0.17 – 0.90) | 0.028 | 0.43 (0.21 – 0.96) | 0.032 |
| Challenges in taking ART (no) | 0.11 (0.01 – 0.93) | 0.042 | 0.12 (0.03 – 0.72) | 0.018 |
| Clinical staff support (no) | 2.87 (1.06 – 7.79) | 0.016 | 3.01 (1.34 – 7.69) | 0.046 |
| Sure, I took ART as directed (yes) | 2.63 (0.83 – 8.32) | 0.104 | 2,65 (1.11 – 7.83) | 0.046 |
| Friend support for Clinic visits (no) | 0.34 (0.12 – 0.94) | 0.037 | 0.31 (0.09 – 0.89) | 0.001 |

#### Age (continuous)

Younger participants (15–19 years) were less likely to achieve viral suppression compared to older participants (20–24 years) after adjusting for other factors. This finding was consistent with the univariable analysis, where age was a significant predictor.

#### Sex

Female participants were more likely to achieve viral suppression than male participants (AOR = 0.43, p-value = 0.032). This shows potential gender disparities in treatment adherence or other contextual factors influencing viral suppression.

#### Challenges in taking ART

Participants who reported not having challenges in taking ART during the preceding three months were significantly less likely to achieve viral suppression (AOR = 0.12, p-value = 0.018). This aligns with the univariable analysis and underscores the critical role of adherence in viral suppression. **Clinical staff support**: Participants who perceived clinical staff as not supportive were significantly more likely to not achieve viral suppression (AOR = 3.01, p = 0.046). This emphasizes the importance of a supportive healthcare environment in improving treatment outcomes.

#### Sure, I took ART as directed

Participants who were not sure they took their ART as directed by health care workers were more likely to not achieve viral suppression (AOR 2,65, p-value = 0.046). After adjusting for confounding variables, this variable became statistically significant, after initially being insignificant

### Qualitative Themes

- **Age-related challenges:** Transitioning from childhood dependence to self-management poses adherence challenges compounded by hormonal and psychosocial changes.
- **Gender dynamics:** Female peers reported higher mutual support; males experienced stigma within peer groups.
- **Educational and work environment:** Boarding schools and work schedules interfered with medication adherence.
- **Family and social support:** Strong family involvement promotes adherence; orphaned youth or those in alternative care faced significant support deficits.
- **Stigma and discrimination:** Both at family and community levels, stigma impairs adherence and clinic engagement.
- **Clinic environment:** Supportive healthcare staff enhance adherence; negative attitudes deter engagement.

## Discussion

This study identified age, sex, ART adherence challenges, clinical staff support and treatment self-efficacy as important correlates of viral load suppression among adolescents and young adults receiving ART in Bulawayo, Zimbabwe.

Older youths aged 20–24 years in our cohort achieved better viral suppression than adolescents aged 15–19 years, suggesting that younger adolescents experience greater difficulty sustaining adherence as they transition from caregiver-supported to self-managed care. This age-related pattern is consistent with regional evidence showing that adolescents transitioning to adulthood often have poorer adherence and lower viral suppression than older age groups, driven by developmental and psychosocial challenges that complicate long-term treatment taking. Reviews of youth living with HIV in sub-Saharan Africa similarly highlight that mid- and late adolescence is a period marked by increased autonomy, evolving identity, stigma, and competing social demands, all of which can disrupt adherence and virological control unless programmes provide developmentally tailored support [9]. These findings reinforce calls for adolescent-centred models of care that specifically support younger adolescents through the transition to greater self-management in order to improve and sustain viral suppression in this age group.

Female participants in this study were more likely to be virally suppressed than males. This pattern is consistent with findings from Zimbabwe, where male sex has been associated with virological non-suppression among adolescents living with HIV [10]. The observed sex difference may be driven by gendered disparities in stigma, disclosure practices, social support and health-seeking behaviour, which influence adherence and sustained engagement in care. Similar gender-related gaps in HIV service outcomes have been reported elsewhere in the region [11], underscoring the importance of sex-responsive adherence and psychosocial support strategies, particularly for young men.

Adherence challenges emerged as a major determinant of viral suppression. Participants who reported no difficulties taking ART were more likely to achieve viral suppression, confirming that adherence remains central to virological control [12], [13]. In our study, adolescents repeatedly described school timetables, fear of disclosure and anticipated stigma as key barriers to both ART adherence and clinic attendance. This is consistent with qualitative work from Western Kenya, where inflexible school schedules and examination timetables were found to clash with ART dosing and clinic appointments, forcing adolescents to choose between attending school and keeping HIV care visits [14]. Similar life-course research among youth in rural Kenya and Uganda has shown that school demands, HIV-related stigma and fears of serostatus disclosure constrain adolescents’ ability to maintain optimal adherence and remain engaged in care [15]. Evidence from Zimbabwean youth further highlights that clinic opening times that are incompatible with school attendance and persistent stigma in health and community settings undermine retention along the HIV care cascade [16]. Taken together, these studies support our participants’ accounts that educational schedules, coupled with fear of disclosure and stigma, remain central structural and social barriers to sustained ART adherence and clinic attendance among adolescents living with HIV in this context.

Beyond schooling, some adolescents described work commitments as a major barrier to adherence, particularly when jobs required travel outside Bulawayo or long shifts that made clinic attendance difficult and left them reluctant to request time off from unsupportive or uninformed employers. Similar challenges with managing treatment around employment and mobility, and the fear of disclosing HIV status in work settings, have been documented in broader sub-Saharan African literature on ART adherence [4]. For participants who had not disclosed their status to bosses, taking medication during work hours was especially challenging, reinforcing evidence that non-disclosure in stigmatizing environments constrains adolescents’ ability to adhere to ART [17]. Adolescents living in alternative care or with relatives who did not know their HIV status described particular difficulties in maintaining treatment, reflecting evidence that non-disclosure, limited family awareness, strained household relationships and mistreatment by non-biological caregivers undermine support for adherence and contribute to missed doses and treatment default [18], [19].

Our finding that adolescents who reported stronger clinical staff support were more likely to achieve viral suppression aligns with prior work showing that good relationships with health providers are among the most commonly cited facilitators of ART adherence, and with programme data from Harare and elsewhere indicating that structured adherence counselling delivered by engaged providers can significantly improve viral suppression among patients[20], [21], [22], [23]. These data reinforce calls for adolescent-friendly services that prioritise relational continuity, empathic communication and repeated adherence counselling contacts as core components of HIV care. In Zimbabwe, the Zvandiri model has shown that peer-delivered community support can reduce virological failure or death among adolescents, underscoring the value of combining clinical care with psychosocial support [24].

Tretament self-efficacy also emerged as an important correlate of viral suppression: young people who felt confident taking ART as prescribed more likely to achieve undetectable viral loads. This is consistent with evidence from adolescent and adult populations showing that stronger ART adherence self-efficacy is associated with better self-reported adherence and higher rates of viral load suppression [25], [26], [27]. Limited treatment literacy also appeared to be an important barrier: some participants were unable to recall or interpret their viral load results, suggesting gaps in understanding of viral load monitoring and its implications for their health, which mirrors recent work from Zimbabwe and the region highlighting poor viral load literacy among adolescents and its potential to undermine adherence and self-management.[28], [29] Taken together with trial evidence that counselling and educational interventions tailored to patients’ literacy levels can improve adherence and virological outcomes, our findings suggest that strengthening counselling on viral load monitoring and ART self-management—particularly by building adolescents’ treatment literacy and confidence—may contribute to improved virological outcomes in this age group.

Economic constraints including transport costs, unstable employment and food insecurity also emerged as important barriers to maintaining treatment and viral suppression. This is consistent with evidence from sub-Saharan Africa showing that poverty, high transportation costs and competing livelihood demands impede clinic attendance and daily pill-taking, and that food insecurity is associated with increased risk of ART non-adherence and incomplete viral load suppression in both African and international cohorts. Longitudinal and cohort studies among adolescents and adults further demonstrate that improved economic well-being is linked to better adherence and viral suppression, underscoring the importance of addressing structural poverty within HIV care programmes [4], [30], [31], [32].

Taken together, our findings suggest that viral suppression among adolescents and young adults in Bulawayo is shaped by an interplay of developmental stage, gender, adherence barriers, clinical support and treatment self-efficacy, rather than by any single factor. This multidimensional pattern is consistent with studies from South Africa, Tanzania and Namibia showing that adolescent viral suppression is influenced by age and gender, structural barriers such as school and transport, and the availability of psychosocial and clinical support. Evidence from Zimbabwe and the region further indicates that differentiated, adolescent-centred models of care—including community-based youth services and peer-led interventions—can improve adherence, retention and viral suppression by addressing school and work-related constraints, strengthening provider–patient relationships and enhancing self-management skills [11], [23], [27], [33]. In line with this literature, our results support the need for differentiated, adolescent-centred interventions in Bulawayo that explicitly tackle school and employment barriers, invest in supportive clinical staff relationships and build treatment literacy and self-efficacy in order to sustain viral suppression in this population.

Finally, the inability of some adolescents and young adults to recall their viral load status highlights an important gap in treatment literacy and patient engagement within this population. Not knowing or remembering one’s viral load may signal ineffective communication between healthcare providers and patients and weaken understanding of the purpose and importance of viral load monitoring for HIV management. This reduces adolescents’ capacity to participate actively in their care and may lower motivation for strict adherence to ART and timely clinic attendance, potentially contributing to suboptimal viral suppression. These observations underscore the need for enhanced, developmentally appropriate education on viral load and treatment goals, so that adolescents understand, remember and can act on their viral load information, thereby supporting improved individual outcomes and progress towards HIV control targets. [29]

### Study Limitations

A few limitations should be considered when interpreting these findings. First, the cross-sectional design precludes causal inference between the psychosocial, structural and clinical factors identified and viral suppression, in line with similar adolescent viral load analyses in the region. Second, viral load results, adherence and other behavioural variables were self-reported and therefore subject to recall and social desirability bias, a common challenge in adolescent HIV research hence may have introduced information bias and limited our ability to fully characterise predictors of viral suppression and non-suppression. Finally, because participants were recruited from one health facility in Bulawayo, our results may not be generalisable to adolescents and young adults who are not engaged in care or accessing other types of services.

### Recommendations and Policy Implications Recommendations

- **Enhance Adolescent and Youth-Friendly Services:** Strengthen the provision of youth-centered HIV care services at Mpilo Centre of Excellence and other facilities by training clinical staff in supportive communication, confidentiality, and addressing unique challenges faced by youths aged 15-24. A positive clinical environment was strongly associated with better viral suppression in this study.
- **Strengthen Treatment Adherence Support:** Develop targeted adherence interventions addressing age-specific barriers, such as transitioning from caregiver-managed to self-managed ART among younger adolescents. Peer support programs like Community Adolescent Treatment Supporters (CATS) should be expanded and adapted to address gender-specific needs, given females showed better viral suppression.
- **Improve Treatment Literacy and Patient Engagement:** Implement enhanced counseling strategies and educational programs that improve youths’ understanding of viral load monitoring, reinforce the importance of consistent ART adherence, and empower patients to remember and engage with their viral load results. This will foster greater self-efficacy and retention in care.
- **Address Socioeconomic Barriers:** Policymakers and program managers should consider interventions that mitigate economic constraints impacting retention and adherence, such as transport subsidies, flexible clinic hours, and integration of HIV services with social support programs addressing food insecurity and employment challenges.
- **Strengthen Psychosocial Support:** Scale up stigma reduction initiatives and ensure access to mental health counseling for adolescents and young adults living with HIV. Enlisting community and family support can enhance adherence and engagement in care.

### Policy Implications

- **Incorporate Youth-Specific Indicators in Monitoring:** National HIV programs should prioritize the routine capture and reporting of retention and viral suppression data disaggregated by narrower youth age bands to identify and address gaps early, reflecting the heterogeneous needs within the 15-24 age group.
- **Resource Allocation for Youth Interventions:** Allocate dedicated funding to sustain differentiated service delivery models tailored for adolescents and young adults that integrate clinical, psychosocial, and socioeconomic support components shown to improve outcomes in this population.
- **Capacity Building for Healthcare Workers:** Institutionalize continuous training on adolescent-friendly care approaches within health systems, focusing on enhancing provider-patient relationships which critically influence viral suppression outcomes.
- **Implement Multi-sectoral Approaches:** Strengthen partnerships between health, education, social welfare, and community sectors to holistically address the multifaceted factors influencing poor retention and viral load suppression among youths.
- **Policy Review and Revision:** Revisit national HIV treatment guidelines to embed specific recommendations on managing adolescent and young adult HIV care challenges, informed by emerging evidence such as this study’s findings from Mpilo Centre of Excellence.

By implementing these recommendations and policy actions, the health system can make significant strides toward achieving UNAIDS 95-95-95 targets among young people, ultimately improving individual health outcomes and reducing HIV transmission in Zimbabwe and similar high-burden settings.

### Recommendations for Future Research

Future research should further explore longitudinal trajectories of adherence and viral suppression from early to late adolescence, including transitions from caregiver-supported to self-managed care, to clarify causal pathways between psychosocial factors, service delivery and virological outcomes. Given the observed importance of treatment self-efficacy, viral load literacy and economic constraints, future work should test scalable packages that combine enhanced VL-focused counselling, mental health support and economic or social protection interventions, particularly among younger adolescents and young men who remain at highest risk of non-suppression.

## Conclusion

This study highlights critical factors influencing poor retention to care and viral load suppression among youths aged 15 to 24 years at Mpilo Centre of Excellence, Zimbabwe. Younger age, male sex, challenges in ART adherence, perceived lack of clinical staff support, and low treatment self-efficacy were significantly associated with suboptimal viral suppression. The findings emphasize the complex interplay of demographic, behavioral, clinical, and psychosocial determinants shaping HIV treatment outcomes in this vulnerable population. Improving viral suppression and retention among adolescents and young adults requires multifaceted interventions that address both individual adherence barriers and systemic factors such as healthcare provider attitudes and socioeconomic constraints. Enhanced youth-friendly services, intensified adherence support, and strengthened patient education to improve treatment literacy and engagement are imperative. These efforts are vital to advancing Zimbabwe’s progress towards UNAIDS 95-95-95 targets and reducing HIV transmission in this age group. Future research and programmatic strategies should prioritize tailored approaches that acknowledge the unique developmental and contextual needs of young people living with HIV.

## Data Availability

Quantitative data are within the manuscript and all qualitative data can be availed.

## Acknowledgements

We would like to express our sincere gratitude to all individuals and institutions who contributed to the successful completion of this study. We are especially indebted to the study participants, who generously gave their time and shared their experiences, making this work possible. We also thank the data collectors, whose commitment in the field and meticulous attention to data quality greatly strengthened this work.

We are deeply grateful to the staff members of the National AIDS Council, Mpilo Centre of Excellence, AIDS Health Foundation, and Million Memory Project Zimbabwe for their invaluable support with coordination, mobilisation, and logistical arrangements throughout the study. Their technical input and operational assistance were instrumental at every stage, from protocol development to implementation and data management.

This work was funded by the National AIDS Council. We further acknowledge the constructive feedback provided by colleagues and mentors during data analysis and manuscript drafting, and we appreciate the editorial and peer review processes that will help refine and improve this manuscript prior to publication.

## References

[1] Gwanzura Clorata, Matare Takura, Apollo Tsitsi, and Ministry of Health and Child Care, “Taking Differentiated Service Delivery to Scale in Zimbabwe: Championing Data for Decision-Making,” Nov. 2019.

[2] C. Jimu, K. Govender, R. Kanyemba, and M. J. O. Ngbesso, “Experiences of intimate relationships, stigma, social support and treatment adherence among HIV-positive adolescents in Chiredzi district, Zimbabwe,” African Journal of AIDS Research, vol. 20, no. 3, 2021, doi: 10.2989/16085906.2021.1979059.

[3] I. Yigit et al., “Longitudinal Associations of Experienced and Perceived Community Stigma With Antiretroviral Therapy Adherence and Viral Suppression in New-to-Care People With HIV: Mediating Roles of Internalized Stigma and Depression Symptoms,” J. Acquir. Immune Defic. Syndr. (1988)., vol. 95, no. 3, 2024, doi: 10.1097/QAI.0000000000003360.

[4] J. Magura, S. R. Nhari, and T. I. Nzimakwe, “Barriers to ART adherence in sub-Saharan Africa: a scoping review toward achieving UNAIDS 95-95-95 targets,” 2025. doi: 10.3389/fpubh.2025.1609743.

[5] P. Ondoa et al., “Access to HIV Viral Load Testing and Antiretroviral Therapy Switch Practices: A Multicountry Prospective Cohort Study in Sub-Saharan Africa,” AIDS Res. Hum. Retroviruses, vol. 36, no. 11, 2020, doi: 10.1089/aid.2020.0049.

[6] S. Moyo et al., “Children and adolescents on anti-retroviral therapy in Bulawayo, Zimbabwe: How many are virally suppressed by month six?,” F1000Res., vol. 9, 2020, doi: 10.12688/f1000research.22744.1.

[7] W. M. Han et al., “Global estimates of viral suppression in children and adolescents and adults on antiretroviral therapy adjusted for missing viral load measurements: a multiregional, retrospective cohort study in 31 countries,” Lancet HIV, vol. 8, no. 12, 2021, doi: 10.1016/S2352-3018(21)00265-4.

[8] “Zimbabwe National HIV Estimates (2023),” 2023.

[9] K. Moomba, T. Crowley, and B. van Wyk, “Tracking viral control in adolescents on antiretroviral therapy in Lusaka, Zambia: A retrospective cohort analysis,” South. Afr. J. HIV Med., vol. 26, no. 1, 2025, doi: 10.4102/sajhivmed.v26i1.1665.

[10] V. Simms et al., “Risk factors for HIV virological non-suppression among adolescents with common mental disorder symptoms in Zimbabwe: a cross-sectional study,” J. Int. AIDS Soc., vol. 24, no. 8, 2021, doi: 10.1002/jia2.25773.

[11] E. F. Okonji, B. van Wyk, F. C. Mukumbang, and G. D. Hughes, “Determinants of viral suppression among adolescents on antiretroviral treatment in Ehlanzeni district, South Africa: a cross-sectional analysis,” AIDS Res. Ther., vol. 18, no. 1, 2021, doi: 10.1186/s12981-021-00391-7.

[12] G. J. Mollel et al., “Viral suppression and associated factors among adolescents living with HIV in rural Tanzania,” HIV Med, vol. 22, no. SUPPL 3, pp. 87–88, 2021, [Online]. Available: https://www.embase.com/search/results?subaction=viewrecord&id=L636553133&from=export

[13] Z. Sithole et al., “Virological failure among adolescents on ART, Harare City, 2017- a case-control study,” BMC Infect. Dis., vol. 18, no. 1, 2018, doi: 10.1186/s12879-018-3372-6.

[14] L. Wiggins et al., “‘They can stigmatize you’: a qualitative assessment of the influence of school factors on engagement in care and medication adherence among adolescents with HIV in Western Kenya,” Health Educ. Res., vol. 37, no. 5, 2022, doi: 10.1093/her/cyac018.

[15] J. Johnson-Peretz et al., “‘I was still very young’: agency, stigma and HIV care strategies at school, baseline results of a qualitative study among youth in rural Kenya and Uganda,” J. Int. AIDS Soc., vol. 25, no. S1, 2022, doi: 10.1002/jia2.25919.

[16] C. D. Chikwari et al., “Differentiated care for youth in Zimbabwe: Outcomes across the HIV care cascade,” PLOS Global Public Health, vol. 4, no. 2 February, 2024, doi: 10.1371/journal.pgph.0002553.

[17] R. Khan, E. C. Garman, and K. Sorsdahl, “Perspectives on Self-Disclosure of HIV Status among HIV-Infected Adolescents in Harare, Zimbabwe: A Qualitative Study,” J. Child Fam. Stud., vol. 32, no. 12, 2023, doi: 10.1007/s10826-023-02612-1.

[18] J. Busza, E. Dauya, T. Bandason, H. Mujuru, and R. A. Ferrand, “‘I don’t want financial support but verbal support.’ How do caregivers manage children’s access to and retention in HIV care in urban Zimbabwe?,” J. Int. AIDS Soc., vol. 17, 2014, doi: 10.7448/IAS.17.1.18839.

[19] B. E. van Wyk and L. A. C. Davids, “Challenges to HIV treatment adherence amongst adolescents in a low socio-economic setting in Cape Town,” South. Afr. J. HIV Med., vol. 20, no. 1, 2019, doi: 10.4102/sajhivmed.v20i1.1002.

[20] N. Croome, M. Ahluwalia, L. D. Hughes, and M. Abas, “Patient-reported barriers and facilitators to antiretroviral adherence in sub-Saharan Africa,” 2017. doi: 10.1097/QAD.0000000000001416.

[21] E. Moyo, P. Moyo, H. Mangwana, G. Murewanhema, and T. Dzinamarira, “Facilitators and Barriers to Antiretroviral Therapy Adherence Among Adolescents and Young Adults in Sub-Saharan Africa: A Scoping Review,” 2025. doi: 10.3390/adolescents5020010.

[22] O. Mukuku, K. Govender, and S. O. Wembonyama, “Barriers and facilitators to HIV viral load suppression among adolescents living with HIV in Lubumbashi, Democratic Republic of the Congo: A qualitative study,” PLoS One, vol. 20, no. 3 March, 2025, doi: 10.1371/journal.pone.0320417.

[23] F. K. Munyayi and B. van Wyk, “Experiences of support by unsuppressed adolescents living with HIV and their caregivers in Windhoek, Namibia: a qualitative study,” Front. Public Health, vol. 12, 2024, doi: 10.3389/fpubh.2024.1380027.

[24] W. Mavhu et al., “Effect of a differentiated service delivery model on virological failure in adolescents with HIV in Zimbabwe (Zvandiri): a cluster-randomised controlled trial,” Lancet Glob. Health, vol. 8, no. 2, 2020, doi: 10.1016/S2214-109X(19)30526-1.

[25] N. Gitahi, S. Wahome, E. A. Bukusi, and P. Memiah, “The role of self-efficacy in HIV treatment adherence and its interaction with psychosocial factors among HIV positive adolescents in transition to adult care in Kenya,” Vulnerable Child. Youth Stud., vol. 17, no. 4, 2022, doi: 10.1080/17450128.2021.1954736.

[26] S. Ashaba et al., “Correlates of HIV treatment adherence self-efficacy among adolescents and young adults living with HIV in southwestern Uganda,” PLOS Global Public Health, vol. 4, no. 9, 2024, doi: 10.1371/journal.pgph.0003600.

[27] S. Mabizela and B. van Wyk, “Viral suppression among adolescents on HIV treatment in the Sedibeng District, Gauteng province,” Curationis, vol. 45, no. 1, 2022, doi: 10.4102/curationis.v45i1.2312.

[28] S. C. Kalichman et al., “Randomized clinical trial of HIV treatment adherence counseling interventions for people living with HIV and limited health literacy,” J. Acquir. Immune Defic. Syndr. (1988)., vol. 63, no. 1, 2013, doi: 10.1097/QAI.0b013e318286ce49.

[29] S. Bernays, J. Lariat, F. Cowan, B. Senzanje, N. Willis, and Z. M. Nenguke, “‘They test my blood to know how much blood is in my body’: the untapped potential of promoting viral load literacy to support adherence and viral suppression among adolescents living with HIV,” J. Int. AIDS Soc., vol. 26, no. 10, 2023, doi: 10.1002/jia2.26153.

[30] P. M. Musumari et al., “Food insecurity is associated with increased risk of non-adherence to antiretroviral therapy among HIV-infected adults in the Democratic Republic of Congo: A cross-sectional study,” PLoS One, vol. 9, no. 1, 2014, doi: 10.1371/journal.pone.0085327.

[31] D. M. Tuller, D. R. Bangsberg, J. Senkungu, N. C. Ware, N. Emenyonu, and S. D. Weiser, “Transportation costs impede sustained adherence and access to HAART in a clinic population in Southwestern Uganda: A qualitative study,” AIDS Behav., vol. 14, no. 4, 2010, doi: 10.1007/s10461-009-9533-2.

[32] J. I. Steinert, Y. Shenderovich, M. Smith, S. Zhou, E. Toska, and L. Cluver, “Economic Well-being and Associated Mediating Pathways to Improved Antiretroviral Therapy Adherence Among Adolescents Living With HIV: A Prospective Cohort Study in South Africa,” J. Acquir. Immune Defic. Syndr. (1988)., vol. 91, no. 4, 2022, doi: 10.1097/QAI.0000000000003071.

[33] A. Asantiel et al., “Viral suppression and adherence in adolescents living with HIV in rural Tanzania,” PLoS One, vol. 19, no. 12 December, 2024, doi: 10.1371/journal.pone.0315866.

